# “Average of neck circumference in Latin American adults: protocol for a systematic review and meta-analysis”

**DOI:** 10.1101/2020.09.18.20197442

**Authors:** Patricia Espinoza Lopez, Kelly Fernández Landeo, Rodrigo Pérez Silva Mercado, Jesús Quiñones Ardela, Rodrigo Carillo Larco

## Abstract

**Background:** Because these are inexpensive and (fairly) easy to conduct, anthropometric measurements have been widely used as risk factors for many diseases (e.g., cardiovascular or cancer). Recently, there have been several reports pinpointing the association between neck circumference and obstructive sleep apnea, cardiovascular and metabolic diseases. These conditions are highly prevalent Latin American and the Caribbean (LAC), where they carry a large disease and disability burden. Neck circumference arises as a potential complement of established anthropometric measurements, namely weight, height and waist/hip circumference. However, unlike these well-known measurements, evidence about neck circumference is scarce and has not been systematically collected in LAC.

**Objective:** To summarize the average neck circumference in LAC adults; and, exploratorily, to summarize the prevalence of large neck circumference (i.e., neck/cervical obesity).

**Methods:** Systematic review and meta-analysis. We will conduct a search in OVID (MEDLINE, Embase, Global Health) and LILACS. Search terms include those related to neck circumference, along with countries in LAC. We seek observational studies with a random sample of the general population, closed populations (e.g., workers), and patients; results will be presented for each of these groups. We will screen titles and abstracts; we will study in detail the selected papers. Both phases will be conducted by two reviewers independently. We will develop an extraction form to collate: country/year of data collection, methods of data collection, average neck circumference and, if available, prevalence of large neck circumference. Data extraction will be conducted by two reviewers independently. We will use the tool proposed by Hoy et al. and the Newcastle-Ottawa scale to assess the risk of bias.

**Conclusions:** This systematic review and meta-analysis will provide the average neck circumference in LAC adults. Currently, evidence about neck circumference in LAC has not been systematically studied, appraised or summarized. This work will provide information about this novel anthropometric measurement, and spark attention about its role as a potential anthropometric indicator and health risk factor.

**Conflicts of interest:** All authors declare to have no conflicts of interest.

**Funding:** Rodrigo M Carrillo-Larco is supported by a Wellcome Trust International Training Fellowship (214185/Z/18/Z).

## BACKGROUND

Among the adult population in Latin America and the Caribbean (LAC), the prevalence of overweight has consistently been above the global average. [1] In addition, while overweight rates in LAC have almost doubled between 1975 and 2016, obesity practically tripled, moving from 7% to 24%. [1] Overweight and obesity are established risk factors for cardio-metabolic diseases. In recent years, however, large neck circumference (NC) has raised as a strong risk factor for cardio-metabolic and other diseases including obstructive sleep apnea. [3, 4, 7, 10, 11, 12]

NC has been correlated with other anthropometric measurements, which are established cardio-metabolic risk factors, for example: body mass index, waist and hip circumference, and waist-to-hip ratio; though the strength of the correlation may vary by gender, age and ethnicity, which calls to study NC in different populations. [2–5] Furthermore, NC offers advantages over other anthropometric measurements. In comparison to body mass index, NC could better distinguish between central and peripheral obesity; and in comparison to waist circumference, which is a good indicator for central obesity, NC will not change during the day, will not be biased by breathing or by postprandial distention. [6, 7]

NC raises as a novel anthropometric indicator strongly associated with several diseases. As it is the case with other anthropometric indicators, NC is also inexpensive and probably not troublesome to incorporate in epidemiological studies and population-based surveys; nevertheless, WC may have additional strengths over other anthropometric indicators regularly collected (e.g., body mass index). In contrast to other anthropometric indicators, mean levels of NC and prevalence estimates of large NC have not been critically appraised or summarized globally or in LAC, where anthropometric indicators play a relevant role in the identification, risk stratification and management of cardio-metabolic diseases. Consequently, we aim to summarize mean levels of NC and prevalence of large NC (i.e., cervical obesity) in adults in LAC, both in the general population and selected groups.

## METHODS

### Study design

This systematic review and meta-analysis will be conducted according to the preferred reporting items for systematic reviews and meta-analyses (PRISMA guidelines). [13]

### Eligibility criteria

- **Participants:** This study will include adult individuals (18+ years old) either from the general population, captive/closed populations (e.g., workers), or patients in the context of any healthcare facility. We will only include people in countries in LAC.
- **Exposure:** NC measured by any method.
- **Comparison:** Not applicable.
- **Types of studies** We will include observational studies including cross-sectional, case-control and cohort studies.

### Exclusion criteria

- The following study designs will be excluded: case reports, case series, letter to the editor, editorial, narrative review, clinical trials and systematic reviews. If there were any systematic review on the matter, we will revise its references to identify relevant original sources.
- We will exclude the studies in which the neck circumference was not measured directly.
- We will exclude the studies with LAC immigrants in countries outside the LAC region.
- We will exclude the studies with patients who have conditions that could disturb or bias the neck circumference measurement, e.g., cervical masses, thyroid disease, cervical fractures and congenital anomalies.

### Literature Search and Data collection

- We will conduct the search in MEDLINE, Embase and Global Health, these three will be searched through OVID; we will also search LILACS. The complete search strategy can be found in Appendix 1.
- Two authors, independently, will screen titles and abstracts looking for studies that meet the selection criteria. Two reviewers independently will study in detail the full-text of the selected publications. Any disagreement will be solved by consensus or by a third author.
- We will record the reason for exclusion in the full-text phase. We will summarize the number of included and excluded publications following the PRISMA flow diagram.

### Data extraction

We will develop a data extraction form in a spreadsheet, which will be piloted with a random sample of five selected publications; if need be, the extraction form will be updated and will not be modified thereafter. Two reviewers independently will carry out data collection. Disagreements will be resolved by discussion until arriving at a consensus, and if that option fails, by consulting with a third party.

The data extracted will include:

- Study details: First author, corresponding author, article title, country, year of publication and year of data collection.
- Sample size: Number of participants, population source (general population, closed population or patients), characteristics of the study population (sex and age), inclusion criteria and exclusion criteria.
- Study methodology: Method of NC measurement.
- Results: Mean levels of NC and prevalence estimates of large NC.

If we find any publication in which the methodology or results are unclear, we will try to contact the corresponding author. In case we do not have an answer from the corresponding author and doubts could not be solved by other means, the original publication will be excluded.

If data from a study was reported in more than one publication, we will only select the paper with the most complete data for our research purposes (e.g., largest sample size or reporting both mean levels and prevalence estimates).

### Risk of Bias and Methodological Quality of Studies

We will assess the risk of bias by duplicate and independently using different tools according to the design of the study:

- Risk of bias for prevalence studies of Hoy et al [14].
- Risk of bias for cohort and case-control studies following the Newcastle-Ottawa scale [15].

If any discrepancy appears, it will be discussed until reaching a consensus between two reviewers. We will present a table with the results of this assessment.

### Statistical analysis

We will present data from included reports using tables summarizing the main findings. We will also provide a narrative synthesis of the evidence found, and we will present tables with the most important data from the included publications. A quantitative summary will be conducted if there are at least three studies with low heterogeneity. We will pool mean levels and prevalence estimates following a random-effects meta-analysis. We will assess heterogeneity with the I^2^ statistic. [16, 17] If feasible (i.e., enough number of publications), we will assess publication bias visually with funnel plots. We will conduct all statistical analyses using Stata 15.1.

## Data Availability

Not applicable

## Appendix 1 APPENDIX Search Strategy for Ovid Search Form (Medline, EMBASE and Global Health)

1. Neck girth
2. Neck circumference
3. Neck obesity
4. Cervical obesity
5. Cervical circumference
6. Cervical girth
7. 1 or 2 or 3 or 4 or 5 or 6
8. (“Antigua and Barbuda” or “Argentina” or “Aruba” or “Bahamas” or “Barbados” or “Belize” or “Bolivia” or “Brazil” or “United States Virgin Islands” or “British Virgin Islands” or “Cayman Islands” or “Chile” or “Colombia” or “Costa Rica” or “Cuba” or “Curacao” or “Dominica” or “Dominican Republic” or “Ecuador” or “El Salvador” or “Grenada” or “Guatemala” or “Guyana” or “Haiti” or “Honduras” or “Jamaica” or “Mexico” or “Nicaragua” or “Panama” or “Paraguay” or “Peru” or “Puerto Rico” or “Saint Kitts and Nevis” or “Saint Lucia” or “Saint Vincent and the Grenadines”or “Suriname” or “Trinidad and Tobago” or “Turks and Caicos Island” or “Uruguay” or “Venezuela” or “Latin America” or latin amer$ or “South America” or south amer$ or “Central America” or central amer$ or “Caribbean Region”)
9. 7 and 8

## Notes

### Competing Interest Statement

The authors have declared no competing interest.

### Author Declarations

This is a systematic review and meta-analysis protocol. No human subjects were involved in this project. Thus, this study was classified as non-human subject research. Therefore, no approval was needed from an IRB/ethics committee

